# Extrapolation of Infection Data for the CoVid-19 Virus and Estimate of the Pandemic Time Scale

**DOI:** 10.1101/2020.03.26.20044081

**Authors:** Walter Langel

## Abstract

Predictions about the further development of the Corona pandemic are widely diverging. Here, a simple yet powerful algorithm is introduced for extrapolating infection rate and number of total infections from available data. The calculation predicts that under present conditions the infection rate in Germany will culminate in a few weeks and decrease to low values by mid-June 2020. Total number of infections will reach several 100000 though. A refinement of the calculation is presented in the supplemental material and shows that the lock down in Germany has reduced the total number of infections from a target value of 338 000 to 184 000, corresponding to a decrease of about 45%.

## Introduction

News pages like [1] publish data on the CoVid19 infection for several countries as a function of time. The total infection numbers show a dramatic increase, and so far no decrease yet with the exception of China (P.R.C). The increase is in general called “exponential”, which implies that finally a significant part of the population will be infected. However, such an exponential function cannot explain the saturation of infection numbers as observed for China (**Figure 1**).

**Figure 1:**
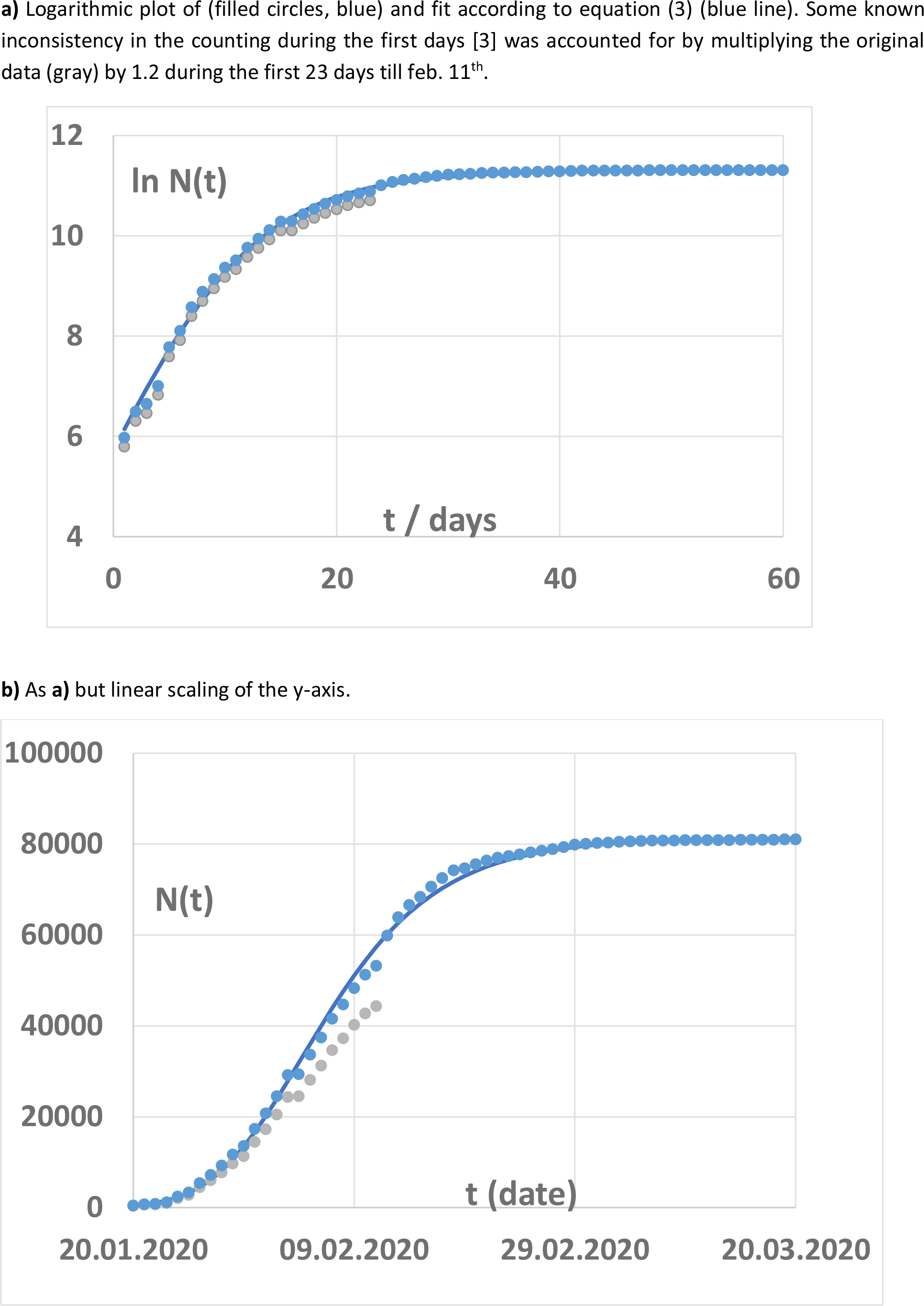
Confirmed infections in China.

Obviously, it is of great interest to have some forecast on the extension of the corona crisis. Many measures taken by governments only extend for weeks so far, but other sources discuss two years of ongoing spreading of the virus and an infection from 10 million to about 2/3 of the total German population [2]. In this worst case, a death rate of e.g. 2% would result in 1.1 millions of victims.

For Germany, the maximum number of infections obviously is by far not yet reached (by march, 23^rd^, 2020). On the other hand a logarithmic plot shows a significant curvature (**Figure 2a**), suggesting that the number of confirmed infection cases already increases significantly less than strictly exponential.

**Figure 2:**
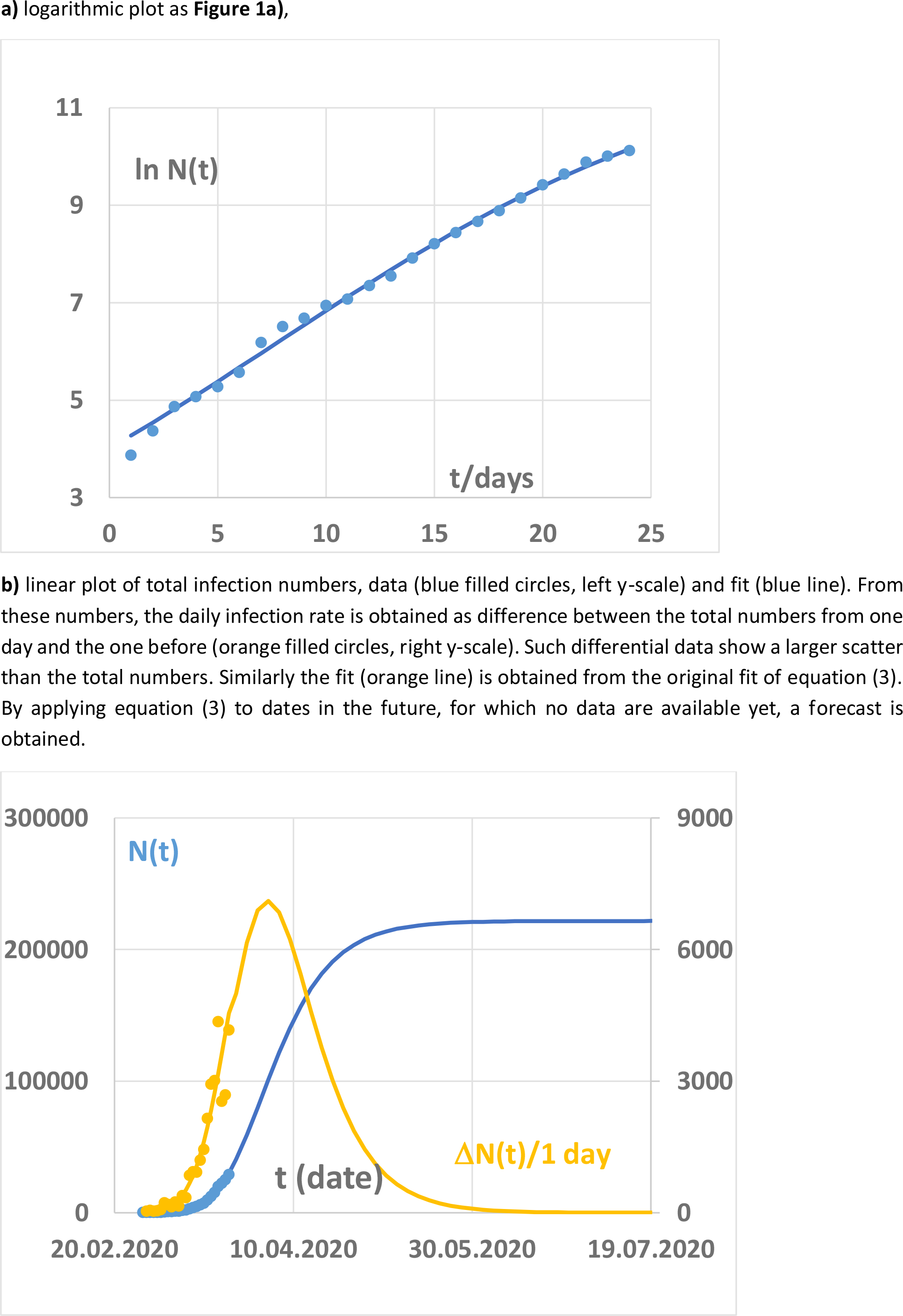
Data for Germany till march, 23^rd^ 2020.

In this communication I try to derive information on the length of the infection period from the deviation of the infection rate from a strict exponential increase. A fit for the total infection numbers as a function of time is presented. This calculation uses an analytical function and is based on three parameters only. The procedure is validated using the data from China, which has overcome the climax of the infection rate already, and then applied to data from Germany and Italy.

## Method

A function for fitting the logarithm of the reported figures of total infection rates is defined by

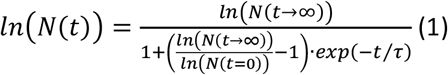

For the fit of equation (1) to the data points, the standard solver in Microsoft Excel 2016 was applied to the logarithm of the number *N*(*t*) of confirmed infections as taken in steps of one day from the website [1], and the least squares error with respect to equation (1) was minimized by varying the three parameters *ln*(*N*(*t* = 0)), *ln*(*N*(*t* → ∞)), and *τ*. Final values are quoted in **Table 1**. The data were weighted by 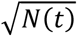to account for poor statistics of small numbers.

**Table 1:** Fitparameter for equation (3). The parameters result in the initial doubling times *τ*_2_ for the total number of CorVid19 infections and the maximum numbers *N*(*t* → ∞) in three countries.

|  | China | Germany | Italy |
| --- | --- | --- | --- |
| $\ln(N(t=0))$ | 5.73 | 4.01 | 2.39 |
| $\ln(N(t \rightarrow \infty))$ | 11.31 | 12.31 | 12.93 |
| $N(t \rightarrow \infty)$ | 81 300 | 220 000 | 410 000 |
| $\tau/d$ | 6.72 | 10.55 | 15.39 |
| $\tau_2/d$ | 1.65 | 2.70 | 5.86 |

The formula correctly reproduces the limiting asymptotic cases:

- At *t* = 0 we obtain consistently

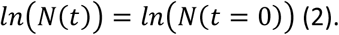 This starting point cannot be taken directly from some scattering initial data but is used as fit parameter. Its actual value depends on the time when counting of cases started.
- For short times, *t* ≪ *τ*, the formula reproduces a nearly exponential increase:

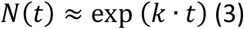

and a time for increasing by a factor of two given

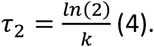 The derivation of equation (3) and of the relation of *k* and *τ*_2_ to the parameters in equation (1) is lengthy and may be found in the appendix. (This main text is, however, fully comprehensible omitting the appendix). For reference purposes, the values of *τ*_2_ are quoted in **Table 1**. The small values of only a few days for this parameter have largely contributed to the fear, which has raised the appearance of CoVid-19.
- The cited data (**Figures 1a, 2a, 3a**) show that already after a few days, significant deviation from this exponential increase has been found in practice. The transition range from initial exponential increase to final saturation is described by the third parameter *τ*, which is characteristic for the rise time of the number of infections.
- At long times, *t* ≫ *τ*, this function attains a maximum of *ln*(*N*(*t* → ∞)) for *t* → ∞. This saturation behavior is generated by the exponential function in the denominator approaching zero for long times *t*.

## Results

The results for the fits are plotted in **Figures 1-3** for China, Germany and Italy, respectively. **Figure 1** shows that the expression (1) with appropriate parameters yields a very good fit of the data for China in the full range from beginning to the end of the pandemic.

For Germany (**Figure 2**), only the beginning of the curve is known so far, and the numbers of confirmed infections are still strongly increasing. Here, the aim of the fit is to predict the maximum number of infections *N*(*t* → ∞), and the time scale, on which this is reached. The orange line suggests that a maximum of more than 7000 new infections per day should be reached early in April, and that the rate should drop to small values by end of May.

The situation in Italy (**Figure 3**) is intermediate, as the fit suggests that the point of maximum infections per day is nearly reached. The maximum infection rate is predicted with more than 9000 per day in early April, and the total number of infections sums up to more than 400 000 in June.

**Figure 3:**
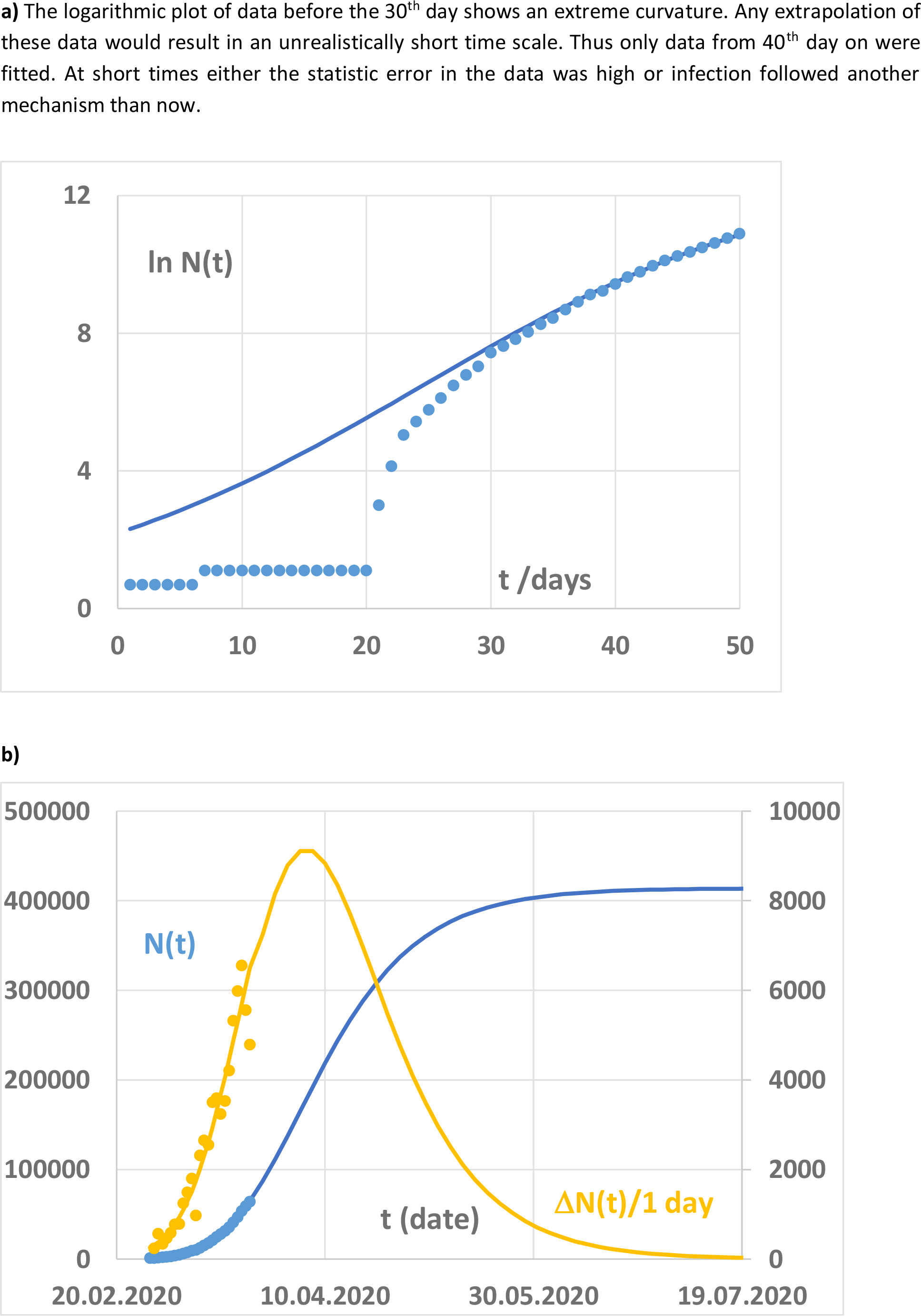
As **Figure 2** for Italy

All three curves suggest that only a small part of the respective population is infected by the spreading of the virus under present conditions and that between 80 000 and 400 000 people may be attained. The period with a high number of new infections should be a few months rather than two years and extend till about June 2020. After this time the further spread of the virus should slow down.

This fit obviously can only refer to the infection under the present conditions with precautions taken so far. It can neither be predicted if more severe political measures reduce the infection rate, or if other mechanisms of virus transfer, which did not yet contribute significantly to the present numbers, may gain importance later.

## Data Availability

All data used were taken from publicly available sources as cited in the manuscript.

## Appendix

### 1. Derivation of the time *τ*_2_ of doubling *N*(*t*) at small times from the fit parameters

With the substitutions

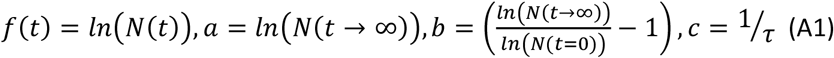

the basic equation (1) is transferred into

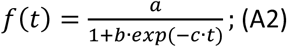

At small times, one may replace

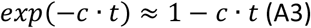

and

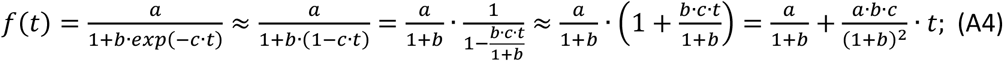

Replacing the substitutions by the original parameters, equation (A5) is obtained:

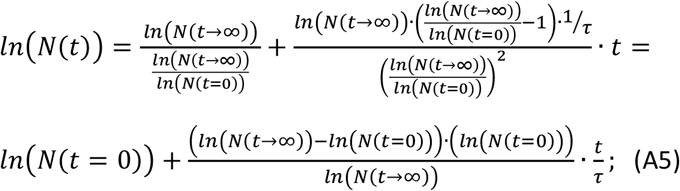

The approximation for small times *t* thus reads

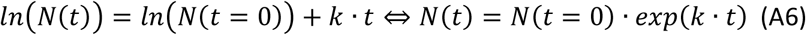

with

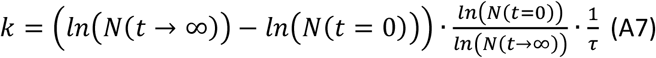

and the time for an increase of the number of infections in the initial state by a factor of two is

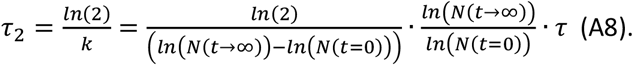

